# Empiric tuberculosis treatment and 12-month mortality among sputum GeneXpert-negative adults living with HIV in Uganda in the era of widespread Antiretroviral therapy: A prospective cohort study

**DOI:** 10.64898/2026.04.04.26350152

**Authors:** Lydia Nakiyingi, Bernard Kikaire, Sarah Nakayenga, Louis Kamulegeya, Elizabeth Nakabugo, Juliet Nkugwa Asio, Bernard Bagaya, Willy Ssengooba, Harriet Mayanja-Kizza, Yukari Manabe

## Abstract

**Background:** In sub-Saharan Africa where both tuberculosis (TB) and HIV are prevalent, empiric TB treatment in people living with HIV (PLHIV) persists due to limited sensitivity of sputum-based TB tests. We evaluated mortality among molecular test-negative presumptive TB adult PLHIV in a population where the majority are or have been on antiretroviral therapy (ART), comparing mortality between those who received empiric TB treatment and those who did not.

**Materials and Methods:** From November 2017 to December 2020, Xpert-negative presumptive TB adult PLHIV were recruited at Mulago Referral Hospital and Kisenyi Health Centre-IV in Kampala, Uganda. Clinical data including TB symptoms, chest X-ray, and empiric TB treatment decision were collected. Laboratory investigations included CD4 cell count, serum cryptococcal antigen (CrAg), urine TB-lipoarabinomannan (TB-LAM), microbiological blood cultures, and sputum mycobacterial growth indicator tube (MGIT) cultures. Participants were followed monthly for 12 months to ascertain vital status.

**Results:** Overall, 300 participants were enrolled; 61.3% inpatients, 55.7% female, median age 37 (IQR 29–45) years, 82.3% on ART, median CD4 206 cells/mm³ (IQR 36–507). Of the 300 participants, 68 (22.7%) received empiric TB treatment, of which 53 (77.9%) were inpatients. 12-month mortality was 31.0% (93/300); 91.4% among inpatients, 72% within three months post-enrolment. Mortality was higher among those who received empiric TB treatment (51.5 vs. 30.2 per 1,000 person-months; p=0.013) compared to those who did not. TB cultures were positive in 5.0% (15/300), of whom seven (46.7%) were also TB-LAM positive. CrAg was positive in 12.3% and 3.7% had positive blood culture.

**Conclusion:** We found high mortality among Xpert-negative PLHIV, particularly those who received empiric TB treatment, despite high ART coverage. Cryptococcal antigenemia and bacteremia were not uncommon. In presence of negative Xpert results in PLHIV, clinicians should perform extensive laboratory evaluations to identify possible comorbidities or alternative non-TB diagnosis.

## Introduction

Tuberculosis (TB) remains the leading cause of death among people living with HIV (PLHIV) globally, accounting for approximately one in every three AIDS-related deaths (1, 2). According to the World Health Organization (WHO) Global Tuberculosis Report 2025 (3), approximately 150,000 HIV/TB co-infected people died in 2024. TB often goes undiagnosed because standard diagnostic tools such as Xpert MTB/RIF (herein referred to as Xpert) and sputum smear microscopy, have limited sensitivity in PLHIV (4, 5). Postmortem studies in sub-Saharan Africa (SSA) report up to 46% of TB cases remain undetected antemortem (6, 7).

Among PLHIV with advanced immunosuppression, clinicians in high-burden HIV/TB settings often resort to empiric TB treatment; initiating TB therapy without microbiological confirmation, including situations where the Xpert result is negative (8, 9). In Uganda, one study found that up to 30% of smear-negative presumptive TB PLHIV received empiric TB treatment (9). The introduction of Xpert MTB/RIF, a more sensitive PCR-based test for MTB detection in HIV/TB high-burden settings, raised expectations for more rapid and accurate TB diagnosis; with decrease in TB-associated morbidity and mortality (10–13). However, a considerable number of PLHIV still test negative on these assays (14, 15), leading to minimal improvements in outcomes. Empiric TB treatment remains common in high HIV/TB burden countries often driven by diagnostic uncertainty and the desire to improve patient outcomes (9, 16–18). While current TB diagnosis and treatment guidelines (19) acknowledge the role of clinical judgement in the diagnosis of TB among PLHIV when microbiological confirmation is unavailable, empiric TB treatment carries the risk of unnecessary exposure to TB treatment, potential undesirable drug effects, and poor patient outcomes (15, 20–22). These challenges are accentuated by concerns of difficulty in monitoring TB treatment response among patients who receive empiric TB treatment as well as managing associated adverse drug reactions (23).

Despite limited data supporting the efficacy of empiric TB therapy (9, 14, 15, 21, 24, 25), clinicians especially in low- and middle-income countries (LMICs) continue to rely on less sensitive diagnostic tools such as smear microscopy, chest X-ray (CXR), response to antibiotics, and clinical presentation to inform and guide their TB treatment decisions particularly among PLHIV (8, 9, 18, 22). Several factors drive empiric TB treatment in LMICs, most of which may not be readily modifiable (9, 22). The low CD4 cell counts commonly observed in this population exponentially increases the risk of TB infection, often tilting the balance toward empiric TB treatment in severely immunocompromised individuals (17, 26). The high rates of incident TB disease and mortality among TB tests-negative PLHIV who do not receive TB treatment after initial evaluation suggest potential survival benefits from empiric TB treatment (27–29). However, randomized controlled trials including REMEMBER (21) and STATIS (25) have shown no mortality benefit from empiric TB treatment, particularly among patients not yet on antiretroviral therapy (ART). These trials were conducted under controlled research conditions with close monitoring. Real-world evidence from routine clinical care in high TB/HIV burden countries, where the majority of patients are on ART, remains limited.

We assessed 12-month mortality among adult sputum Xpert-negative presumptive TB PLHIV in Uganda, comparing those who received empiric TB treatment and those who did not, in a population where the majority are or have been on ART. The aim was to generate real-world data on the outcomes of empiric TB treatment in a high TB/HIV burden setting where ART is readily available. We also investigated non-TB etiologies in this HIV population.

## Materials and Methods

### Study design and setting

This was a prospective study among adult sputum Xpert-negative presumptive TB PLHIV recruited from Kisenyi Health Centre IV (outpatients), a Kampala City Council Authority Clinic and Mulago-Kiruddu National Referral Hospital (MNRH) (inpatients) between 1^st^ November 2017 to 30^th^ December 2020. In Uganda, Xpert was introduced in PLHIV in 2010 at tertiary referral centers and was available as the main TB diagnostic at both facilities during the study period; Xpert Ultra had not yet been introduced. Other investigations for TB at the enrolment sites included chest X-ray (CXR) and smear microscopy. Both sputum TB culture and urine-TB lipoarabinomannan (TB-LAM) for PLHIV were only available for participants in this study.

All TB investigations and treatment at the facilities are free of charge. Patients diagnosed with active TB are treated in accordance with the existing treatment guidelines from the Uganda Ministry of Health National TB program (30).

### Patient recruitment and study population

PLHIV aged 18 years and above, clinically suspected to have active TB as per the WHO guidelines (19) who had undergone routine evaluations for pulmonary TB by the attending team were screened for eligibility. Eligible patients with confirmed HIV, suspected to have active TB based on the WHO criteria, and had received a negative Xpert test were enrolled into the study. Patients who had been initiated on anti-TB medication prior to the enrollment visit were excluded.

Sociodemographic and medical information were obtained from consenting participants including; TB symptoms and signs, previous TB treatment, history of antiretroviral therapy (ART). Information on clinical decision made by the attending clinician regarding empiric TB treatment initiation was obtained from participants’ medical records. Participants provided one spot sputum sample for mycobacterial growth indicator tube (MGIT) TB culture. Blood was collected for microbiological blood cultures, CD4 cell count and serum Cryptococcal Antigen (CrAg) (IMMY CrAg® LFA, Norman, Oklahoma, USA). Each participant provided a urine sample for TB-LAM testing (TB-LAM Alere, Waltham, MA, USA).

CXR findings were obtained from participant records. Participants had monthly telephone follow-ups for 12 months post-enrolment to obtain data on survival status recorded as either dead or alive. For participants who died while in hospital, the date of death (DOD) was recorded immediately, while for those who died after discharge, the DOD was recorded during the telephone follow-up call. However, if the exact DOD was not available, the date of the telephone follow-up was captured as the DOD.

All laboratory procedures were done following standard laboratory procedures at the Mycobacteriology (BSL-3) Laboratory at the Department of Medical Microbiology, Makerere University (College of American Pathologists (CAP: ISO15189) Accredited Laboratory). A participant was considered TB culture positive if MGIT culture was positive, while positive blood culture was defined as the isolation of organisms (bacterial or fungal) from blood culture media. For the TB-LAM test (Alere, Waltham, MA, USA), results were graded from 1+ to 4+.

All study laboratory results were made available to the attending clinicians. Discharged participants were contacted by telephone to deliver results and those whose tests were positive were requested to return for treatment. Participants whose results were positive but could not be contacted by telephone had study home visits performed during which information on any medical treatment, and survival status were obtained.

### Statistical analysis

The primary outcome was survival status at 12 months post-enrolment among presumptive TB PLHIV with negative Xpert test, stratified by empiric TB treatment status. Continuous variables were summarized using medians and inter-quartile ranges (IQR) while categorical variables were summarized using proportions. Using chi-square, we compared the study population characteristics by empiric TB treatment status strata. Kaplan-Meier survival analysis was conducted to estimate time-to-death, and survival probabilities with 95% confidence intervals (95%CI). The log-rank test was used to compare survival curves across different subgroups including empiric TB treatment status and inpatient vs. outpatient setting. Mortality rates were calculated per 1,000 person-months of follow-up, and differences between groups were assessed using mortality rate ratios (RRs) and corresponding 95% CI. Person-time at risk was calculated from the date of enrolment to the earliest time of death, loss to follow-up, or end of the 12-month observation period. The Cox proportional hazards model was used to identify independent predictors of survival. All variables with a p-value < 0.20 in the bivariate analysis were included in the multivariable model. Hazard ratios (HRs) with 95% CI were reported. The assumption of proportional hazards was assessed using the Global test based on the Schoenfeld residuals. All data were analyzed using STATA® version 18.0 (StataCorp, 4905 Lakeway Drive College Station, Texas USA).

### Ethical considerations

The study was approved by the Mulago Hospital Research and Ethics Committee (MHREC# 1120) Kampala, Uganda and the Uganda National Council for Science and Technology (UNCST#HS72ES). All participants provided written informed consent before participation in the study. All consent forms were approved by the above-mentioned ethics committees.

## Results

### Participant characteristics

Overall, 300 Xpert-negative PLHIV participants were eligible for analysis **(Figure 1**); 61.3% (184/300) inpatients, 55.7% (167/300) female with median age 37 (IQR 29-45) years. Majority (82.3%, 247/300) of the participants were taking ART treatment, with median CD4 cell count 206 (IQR 36 - 507) cells/mm^3^. A total of 82/300 (26.7%) participants had been previously treated for drug susceptible TB **(Table 1**). Chest X-ray (CXR) was interpreted as suggestive of TB by the attending clinician in 11 (4.7%) of the participants with CXRs available. Fifteen (5%) participants had positive sputum TB culture, 11 of these were *Mycobacterium tuberculosis* complex (MTBc) while four were *mycobacterium other than tuberculosis* (MOTT). Urine TB-LAM was positive in 23 (7.8%) of the 300 participants, nine (9) of whom had strongly positive LAM bands (2+ and more). Serum CrAg was positive in 37, of whom 3 were also blood culture positive. Blood cultures were positive in 8 additional participants, although 2 were likely contaminated with coagulase-negative staphylococcus **(Table 1)**.

**Figure 1:**
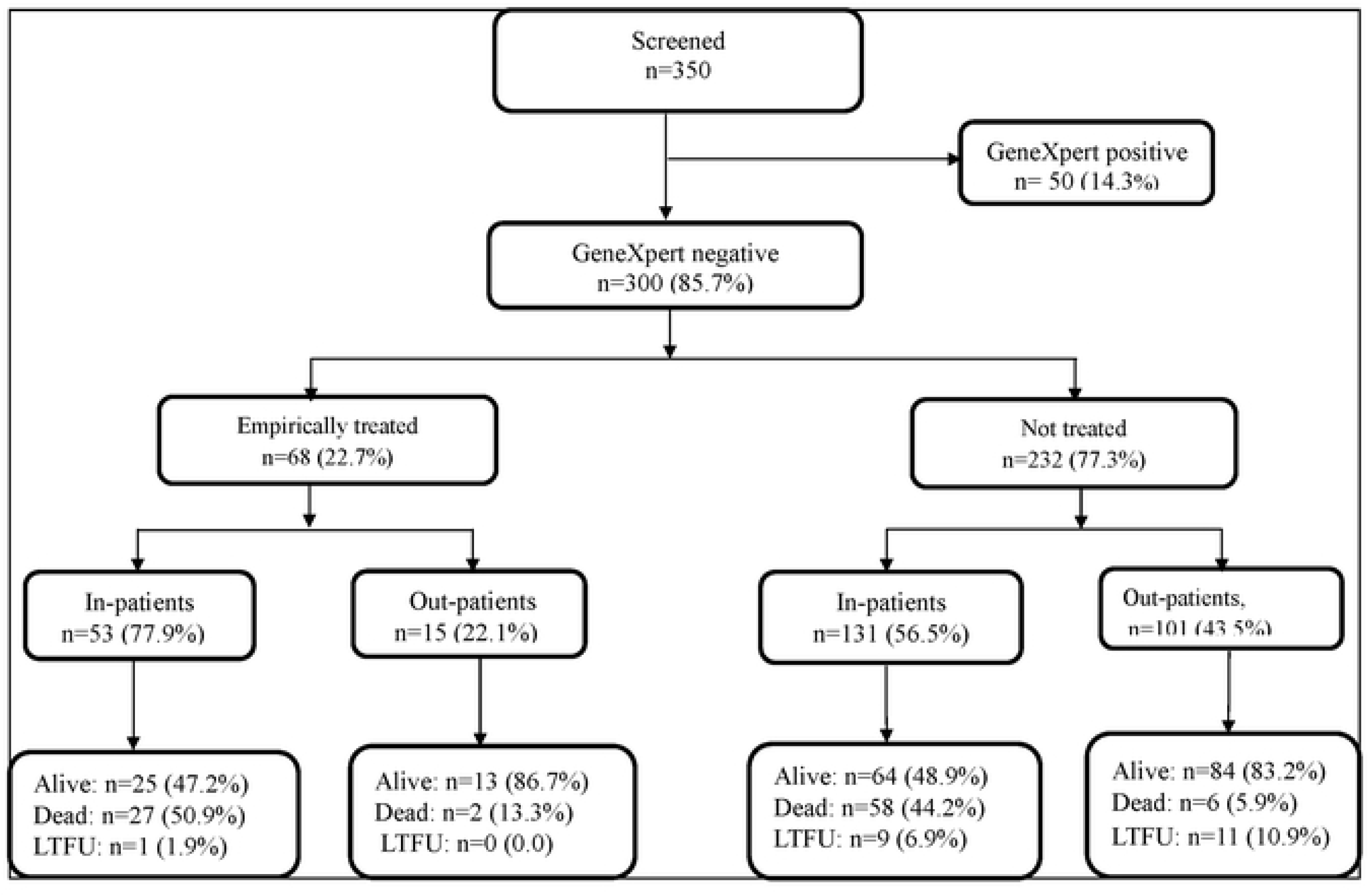
Showing flow of study participants.

**Table 1:** Baseline participant characteristics stratified by empiric TB treatment status.

| Characteristics | Overall<br>(n=300) | Received<br>Empiric TB<br>therapy (n=68) | Not treated for<br>TB (n=232) | p-value |
| --- | --- | --- | --- | --- |
| Median age in years (IQR) | 37 (29-45) | 35 (27-44) | 38 (29-45) | 0.161 |
| Inpatients (hospitalized) | 184 (61.3) | 53 (77.9) | 131 (56.5) | 0.002 |
| Female sex | 167 (55.7) | 35 (51.5) | 132 (56.9) | 0.428 |
| BMI (kg/m<sup>2</sup>, IQR) | 21.2 (18.9-24.9) | 19.9 (18.0-22.5) | 22.2 (18.9-24.9) | 0.002 |
| Fever (Temperature≥37.5°C) | 26 (8.7) | 10 (14.7) | 16 (6.9) | 0.005 |
| Karnofsky score in % (IQR) | 80 (70 - 80) | 70 (70 - 80) | 80 (70 - 80) | 0.003 |
| On ART treatment | 247 (82.3) | 50 (73.5) | 197 (84.9) | 0.030 |
| Duration on ART (median months) | 13 (6 - 36) | 13 (13 - 30) | 13 (5 - 36) | 0.353 |
| Median CD4 count (IQR) | 206 (36-507) | 87 (16-335) | 373 (158-638) | <0.001 |
| Previously received TB treatment | 82 (27.3) | 20 (29.4) | 62 (26.7) | 0.622 |
| CXR findings* (n=234) | 11 (4.7) | 10/68 (14.7) | 1/167 (0.59) | 0.004 |
| Positive sputum TB culture | 15 (5.0) | 11 (16.2) | 04 (1.7) | 0.007 |
| MTB | 11 | 8 | 3 |  |
| MOTT | 04 | 3 | 1 |  |
| Urine TB LAM positive | 23 (7.7) | 16 (23.5) | 07 (3.0) | 0.011 |
| Positive (+) | 14 | 11 | 3 |  |
| Positive (++) | 8 | 5 | 3 |  |
| Positive (+++) | 1 | 0 | 1 |  |
| Positive serum CrAg* n=242 | 37 (15.3) | 33 (24.8) | 4 (3.7) | <0.001 |
| Positive sputum TB culture | 15 (5.0) | 11 (16.1) | 04 (1.7) | <0.001 |
| MTB | 11 | 8 | 3 |  |
| MOTT | 4 | 3 | 1 |  |
| Positive Blood cultures* n=241 | 11 (4.6) | 10 | 1 | - |
| *Cryptococcus Neoformans* | 3 | 3 | 0 |  |
| *Coagulase negative staph* | 2 | 1 | 1 |  |
| *Staphylococcus Aureus* | 1 | 1 | 0 |  |
| *E. coli* | 1 | 1 | 0 |  |
| *Others (Klebsiella oxytoca, Aeromonas, Bacillus, Enterobacter species* | 4 | 4 | 0 |  |

\* means the number of variables is less than the total N=300. These were missing data due to unrecorded data and unperformed study tests.
\*\*CXR read by attending clinician
**Abbreviations:** CrAg, cryptococcal Antigen; TB, tuberculosis; MTB, *Mycobacterium tuberculosis*; MOTT, *Mycobacterial Other than Tuberculosis*; ART, antiretroviral therapy; CXR, chest X-ray; LAM, lipoarabinomannan; IQR, inter-quartile range; BMI, Body Mass Index; E. coli, *Escherichia coli*.

### Empiric TB treatment decision among Xpert-negative PLHIV

Overall, 68 (22.7%) of the 300 study participants received empiric TB treatment, the majority (53/68; 77.9%) coming from the inpatient setting **(Figure 1)**. Compared to those that were not treated, participants who received empiric TB therapy were less likely taking ART at the time of enrolment (73.5% vs. 84.9%; p = 0.030), median CD4 cell counts were significantly lower (87 cells/µL vs. 373 cells/µL; p = 0.001), and had a significantly lower median body mass index (BMI) (19.9 kg/m² vs. 22.2 kg/m²; p = 0.002). Sputum TB culture was positive among 11 (8 MTBc and 3 MOTT) of those who received empiric TB treatment. A comparison of baseline demographic and clinical characteristics by empiric TB treatment decision is presented in **Table 1**.

### 12-months mortality among study participants

At 12 months post-enrolment, of the 300 participants, 31.0% (93/300) had died; 69 (74.2%) of these were on ART, 46.1% (85/184) deaths occurred among inpatients and 6.9% (8/116) among outpatients (p=0.001). The overall total time at risk for the 93 participants who died was 300 person-months, with a mortality rate of 310 deaths per 1000 person-months. A total of 21 (7.0%) participants were lost to follow-up, while 62.0% (186/ 300) were still alive **(Figure 1)**.

Kaplan–Meier survival curves to estimate and compare the cumulative probability of survival over time stratified by key variables including sex, age, empirical TB treatment status, and patient care setting (inpatient vs outpatient) are illustrated in **Figures 2A-2D**. Overall, survival probability declined over the follow-up period, with approximately 31% cumulative mortality by month 12. Most deaths occurred within the first three months **(Figure 2A)**. Survival differed significantly by empiric TB treatment status, patient care setting and body mass index (BMI).

Participants empirically treated for TB had a significantly lower survival compared to those not treated (log-rank p = 0.021), and the decline was more pronounced in the first three months of recruitment, with a gradual decrease in deaths over time **(Figure 2B)**. Similarly, inpatients experienced a significantly lower survival compared to out-patients (log-rank p < 0.001), with cumulative mortality exceeding 60% by month 12 **(Figure 2C)**, and so was underweight when compared to normal body weight (p=0.004) **(Figure 2D)**.

**Figure 2:**
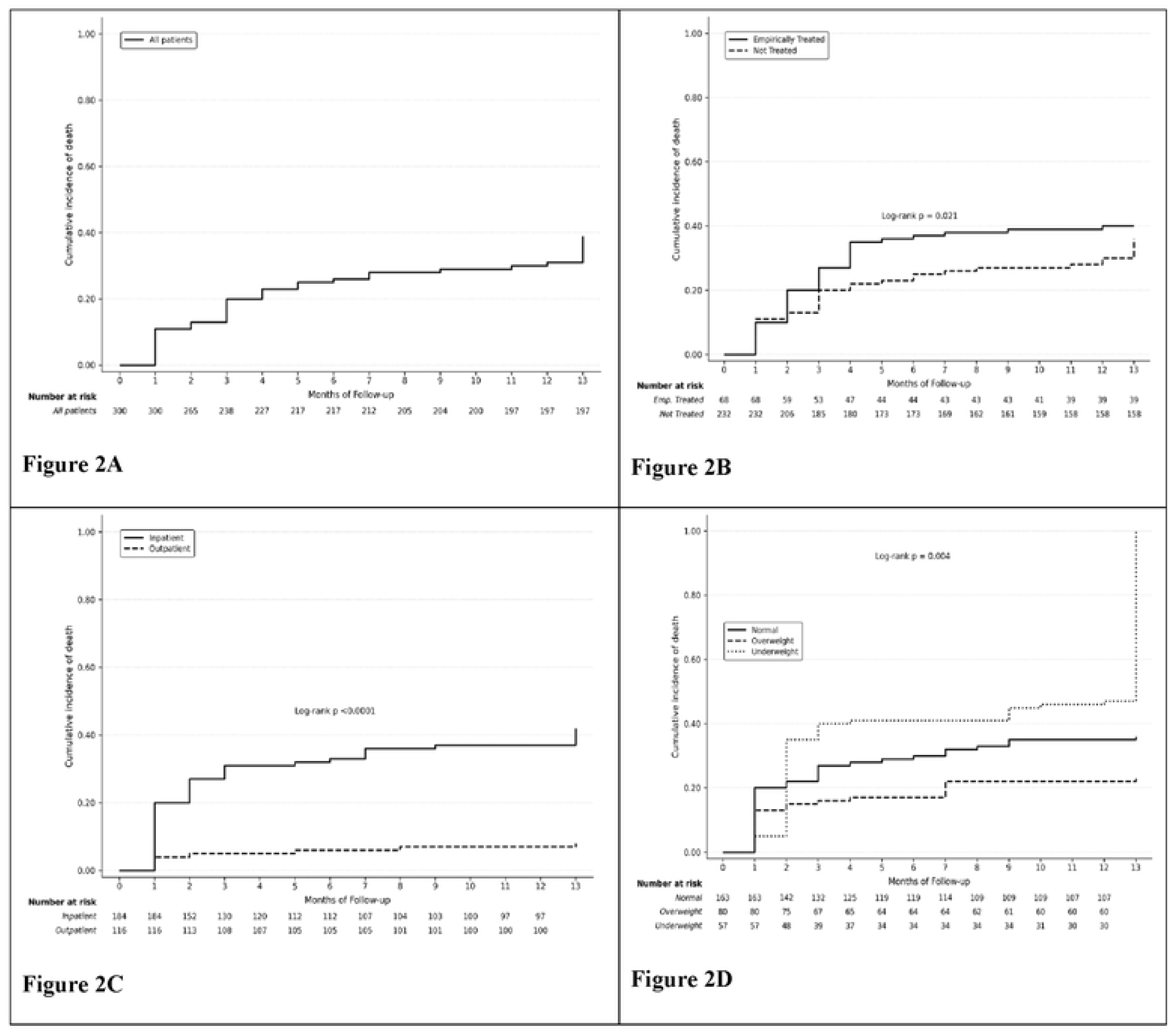
Kaplan–Meier estimates of 12-month mortality among participants. Figure 2A showing Kaplan–Meier 12-month mortality estimates for overall study participants; Figure 2B showing Kaplan–Meier 12-month mortality estimates by empiric TB treatment decision; Figure 2C showing Kaplan–Meier 12-month mortality estimates by care setting (inpatient vs outpatient); Figure 2D showing Kaplan–Meier 12-month mortality estimates by Body Mass Index.

### 12-months mortality and empiric TB treatment decision

Of the 93 participants who died, of whom 69 (74.2%) were taking ART at the time of enrolment, 29/93 (31.2%) had received empiric TB treatment, while 64 (69%) had not. There was a statistically significant difference in proportions of death between those who received empiric TB treatment and those who did not, that is, 42.6% of the 68 empirically treated vs 27.5% of the 232 who were not, log-rank p=0.021. Notably, 7.5% (7) and 8.6% (8) of the 93 deaths had positive TB culture and urine TB-LAM tests respectively, yet in almost all, empiric TB treatment initiation decision had not been made (**Table 2**). A total of 16 (17.2%) of the 93 deaths had positive serum CrAg, of which four had received empiric TB treatment. Cryptococcal treatment was not documented.

**Table 2:** Laboratory results of the 93 participants who died.

|  | Overall deaths,<br>(N=93) | Empiric TB,<br>Treatment, N=26 | Not treated,<br>N=67 |
| --- | --- | --- | --- |
| Positive Urine TB-LAM | 8(8.6) | 2 | 6 |
| Positive MTB Cultures | 07(7.5) | 1 | 6 |
| Positive serum CrAg | 16 (17.2) | 4 | 12 |
| Positive bacterial blood cultures | 7(7.5) | 1 | 6 |

**Abbreviations:** TB, tuberculosis; TB-LAM, lipoarabinomannan; CrAg, cryptococcal Antigen; TB, tuberculosis; MTB, *Mycobacterium tuberculosis*

### Mortality risk among study participants

Mortality risk ratios and mortality rates are presented in **Table 3**. Notably, compared to those not treated, participants empirically treated for TB had a significantly higher mortality rate (51.51, 95% CI: 35.80–74.12 vs 30.15, 95% CI: 23.60–38.52 per 1,000 person-months), as well as a 55% increased risk of death (Risk Ratio, RR; 1.55; 95% CI: 1.09–2.18; p = 0.013). Inpatients had a higher mortality rate (59.78 per 1,000 person-months; 95% CI: 48.33–73.93) compared to outpatients (6.32 per 1,000 person-months; 95% CI: 3.17–12.66), while participants on ART had lower mortality rate compared to those not on ART (29.96, 95% CI 23.66 - 37.93 vs 62.66, 95% CI: 42.00-93.49 per 1,000 person-months) **(Table 3)**.

**Table 3:** Mortality risk and rates for the 300 Xpert-negative PLHIV participants.

| Characteristic | n/N | Mortality Risk Ratio (95%CI) | P-value | Mortality rate per 1000 person-months (95% CI) |
| --- | --- | --- | --- | --- |
| **Sex** |  |  |  |  |
| Male | 47/133 | 1 |  | 41.16 (30.92 - 54.78) |
| Female | 46/167 | 0.78 (0.56 – 1.09) | 0.147 | 29.79 (22.32 - 39.78) |
| **Age (years)** |  |  |  |  |
| < 40 | 53/165 | 1 |  | 36.63 (27.98 - 47.94) |
| ≥ 40 | 40/135 | 0.92 (0.66 – 1.30) | 0.643 | 32.28 (23.68 - 44.01) |
| **Empirically treated for TB** |  |  |  |  |
| No | 64/232 | 1 |  | 30.15 (23.60 – 38.52) |
| Yes | 29/68 | 1.55 (1.09 – 2.18) | 0.013 | 51.51 (35.80 – 74.12) |
| **Patient care setting** |  |  |  |  |
| In-patient | 85/184 | 1 |  | 59.78 (48.33 - 73.93) |
| Out-patient | 8/116 | 0.15 (0.06 – 0.30) | <0.001 | 6.32 (3.17 - 12.66) |
| **On ART** |  |  |  |  |
| No | 24/53 | 1 |  | 62.66 (42.00 - 93.49) |
| Yes | 69/247 | 0.62 (0.43 – 0.88) | 0.008 | 29.96 (23.66 - 37.93) |
| **Previous TB treatment** |  |  |  |  |
| No | 69/211 | 1 |  | 37.66 (29.75 – 47.69) |
| Yes | 22/80 | 0.85 (0.56 – 1.26) | 0.402 | 28.76 (18.94 – 43.68) |
| **Body Mass Index** |  |  |  |  |
| Underweight | 25/57 | 1 |  | 56.43 (38.13 – 83.52) |
| Normal | 54/163 | 0.76 (0.52 - 1.09) | 0.133 | 37.04 (28.37 – 48.36) |
| Overweight/Obese | 14/80 | 0.40 (0.29 – 0.70) | 0.001 | 17.83 (10.56 – 30.11) |
| **Karnofsky score** |  |  |  |  |
| ≥80 | 17/157 | 1 |  | 10.19 (6.33 – 16.38) |
| <80 | 76/143 | 4.92 (3.05 – 7.89) | <0.001 | 74.73 (59.68 – 93.57) |

**Abbreviations:** TB, tuberculosis; ART, antiretroviral therapy

### Factors associated with 12-month mortality among participants

We estimated crude and adjusted mortality rate ratios (MRRs) using Poisson regression models with robust standard errors (**Table 4**). At bivariate, empiric TB treatment (42.7% 95% CI: 32.0 – 55.4, p=0.029), inpatients (46.3%; 95% CI: 39.4-53.9, p=0.001), not taking ART (46.6%, 95% CI: 34.1-61.1, p=0.008), and poor functional status (KPS <80: 52.9%, 95% CI: 44.9-61.3, p=0.001) were associated with higher 12-month mortality in this population. In multivariable Poisson regression, patient care setting (p = 0.008) and functional status (p = 0.001) remained independently associated with mortality. Although participants who received empiric TB treatment had a higher crude mortality (42.7%; 95% CI: 32.0–55.4) than those not treated (27.6%; 95% CI: 22.3–33.9), this difference was not statistically significant after adjusting for other factors (aMRR 1.05; 95% CI: 0.62–1.77; p = 0.850). Other patient characteristics, including ART status, sex, age group,, previous TB treatment, and body mass index, were not significantly associated with mortality in the adjusted model (**Table 4**).

**Table 4:** Factors associated with 12-month mortality among Xpert-negative PLHIV.

| Characteristics |  | Univariate Analysis |  | Multivariate Analysis |  |
| --- | --- | --- | --- | --- | --- |
|  | Mortality rate at 12 months (%) | Crude mortality rate ratio (95% CI) | P-Value | Adjusted mortality rate ratio (95% CI) | P-Value |
| **Sex** |  |  |  |  |  |
| Male | 36.0 (28.5 – 44.9) | 1 |  | 1 |  |
| Female | 27.2 (21.1 – 34.7) | 0.72 (0.47 – 1.12) | 0.15 | 0.79 (0.48 – 1.28) | 0.336 |
| **Age (years)** |  |  |  |  |  |
| < 40 | 32.1 (25.5 – 39.9) | 1 |  | 1 |  |
| ≥ 40 | 29.9 (22.9 – 37.8) | 0.88 (0.57 – 1.37) | 0.577 | 0.82 (0.50 – 1.34) | 0.427 |
| **Empirically treated** |  |  |  |  |  |
| No | 27.6 (22.3 – 33.9) | 1 |  | 1 |  |
| Yes | 42.7 (32.0 – 55.4) | 1.71 (1.05 – 2.76) | 0.029 | 1.05 (0.62 – 1.77) | 0.85 |
| **Patient category** |  |  |  |  |  |
| In-patient | 46.3 (39.4 – 53.9) | 1 |  | 1 |  |
| Out-patient | 6.9 (3.5 – 13.4) | 0.11 (0.50 – 0.22) | <0.001 | **0.26 (0.10 – 0.71)** | **0.008** |
| **On ART** |  |  |  |  |  |
| No | 46.6 (34.1 – 61.1) | 1 |  | 1 |  |
| Yes | 27.9 (22.7 – 34.0) | 0.48 (0.28 – 0.81) | 0.008 | 0.66 (0.36 – 1.19) | 0.167 |
| **Previous TB treatment** |  |  |  |  |  |
| No | 33.3 (27.4 – 40.3) | 1 |  | 1 |  |
| Yes | 26.3 (18.0 – 37.5) | 1.71 (1.06 – 2.76) | 0.029 | 1.11 (0.64 – 1.92) | 0.173 |
| **Body Mass Index** |  |  |  |  |  |
| Underweight | 42.6 (30.9 – 56.6) | 1 |  | 1 |  |
| Normal | 33.4 (26.7 – 41.3) | 0.66 (0.39 – 1.11) | 0.116 | 0.68 (0.39 – 1.18) | 0.173 |
| Overweight/Obese | 17.9 (11.0 – 28.3) | 0.32 (0.16 – 0.64) | 0.001 | 0.56 (0.25 – 1.28) | 0.17 |
| **Karnofsky score** |  |  |  |  |  |
| ≥80 | 11.1 (7.0 – 17.2) | 1 |  | 1 |  |
| <80 | 52.9 (44.9 – 61.3) | 7.34 (4.23 – 12.72) | <0.001 | **3.50 (1.69 – 7.23)** | **0.001** |

277 **Abbreviations:** TB, tuberculosis; ART, antiretroviral therapy

## Discussion

We found high 12-month cumulative mortality among Xpert-negative presumptive TB adult PLHIV despite majority taking ART, mostly occurring in the first three months after enrolment and predominantly among hospitalized participants. Mortality was higher among those who received empiric TB therapy (48.1 per 1,000 person-months) compared to those who did not (30.9 per 1,000 person-months). We also investigated non-TB etiologies in this study population and found that Cryptococcosis and bacteremia were not uncommon. Although empiric TB treatment was initiated in nearly one-in-four Xpert-negative PLHIV, the treatment did not improve survival after adjustment for confounding factors. Inpatient status and low Karnofsky Performance Score (KPS) were independently associated with high mortality.

We report 31% cumulative mortality among Xpert-negative adult PLHIV, despite the majority of participants reporting taking ART and most being virally suppressed. This is consistent with earlier studies that have evaluated mortality in similar populations. In 2017, Heunis et al in South Africa reported a 36.7% mortality among HIV/TB co-infected patients, and 23.8% among those who were smear-negative and had missed early TB diagnosis due to diagnostic challenges (31). The South African study was a 10-year retrospective records review of risk factors for TB mortality in the general population and did not include Xpert-negatives. Similar trends have been observed in Brazil, where late TB diagnosis in PLHIV, particularly those with negative Xpert results, has been associated with delayed treatment and poor outcomes (32). Limited access to advanced diagnostics in resource-limited settings (RLS) like Uganda, and potential misdiagnosis of other pulmonary diseases that mimic TB contribute to poor outcomes (33). A meta-analysis comparing the diagnostic accuracy of tools like Xpert, and the WHO algorithm in smear-negative TB cases revealed that in settings with high HIV/TB prevalence, smear-negative TB cases are often misdiagnosed, leading to delayed treatment and high mortality (34).

Mortality in Xpert-negative adult PLHIV may be due to either missed TB diagnosis or misdiagnosis of non-TB conditions. Studies have shown that smear-negative patients may have other pulmonary or systemic diseases that mimic TB, such as bacterial pneumonia, pneumocystis pneumonia, or malignancies (35). Misdiagnosis or failure to recognize these conditions can delay appropriate treatment and contribute to high mortality. Inadequate diagnostics in RLS further exacerbate these challenges (36, 37). In SSA and other low-income regions, access to comprehensive diagnostic tools, like chest Computed Tomography (CT) scans or bronchoscopy is limited, often resulting in empiric treatment based on clinical suspicion rather than confirmed diagnosis (38).

Our study observed higher mortality rate among patients who received empiric TB treatment. This finding contrasts with the study by Huerga et al. (2019) (8), which reported no association between TB treatment and increased mortality among PLHIV, and another 8-week follow-up study which found reduced risk of mortality at 8 weeks of empiric treatment in a population of smear-negative PLHIV (18). Whereas Huerga et al. evaluated six-month mortality among all patients initiated on TB treatment regardless of whether treatment was empiric or based on bacteriological confirmation, our study was restricted to Xpert-negative PLHIV. This narrower focus may reflect a subgroup with more advanced disease, greater diagnostic uncertainty, and higher baseline mortality risk, potentially accounting for the higher mortality observed in our study. The longer duration of follow-up (up to 12 months) in our study also explains the higher mortality, although majority of our participants died within the first three months.

Nevertheless, our findings are consistent with a study from Brazil, which reported higher mortality among PLHIV with negative sputum smear who received empiric TB treatment (39). Similarly, a study conducted in Uganda found that smear-negative PLHIV initiated on empiric TB treatment experienced higher mortality after one-year of follow-up; however, this association was no longer significant after adjustment for CD4 cell count and antiretroviral therapy (ART) use (40). Our study further supports the earlier clinical trials (21, 24, 25) that reported no benefit of empiric TB treatment in reduction of mortality among ART naïve PLHIV with advanced disease. Importantly, REMEMBER (21) and STATIS (25) trials found that empiric TB treatment did not reduce mortality in adult PLHIV with advanced HIV disease, particularly among patients not yet on ART. Our findings extend these earlier trial results to a real-world setting where the majority of PLHIV are on or have been on ART, suggesting that even with extensive ART coverage, empiric TB treatment may not improve survival.

The high mortality in our study could also be explained by the fact that we had a population of much sicker patients compared to the earlier studies as was shown by the large proportion of inpatients with low CD4 counts and low KPS. Advanced HIV disease and other immune suppressive states are significant contributors to high mortality rate (16, 35). Similar to our study, many Xpert-negative patients are co-infected with HIV and often present with advanced disease marked by low CD4 counts (16, 35). Further, advanced HIV disease makes TB diagnosis challenging since standard tests become less sensitive. Indeed, being inpatient and having low KPS were found significant predictors of mortality in our study. It is also possible that once empiric TB treatment is initiated, clinicians are unlikely to perform further investigations for other possible non-TB conditions leading to high mortality among Xpert-negative PLHIV. In a Ugandan study conducted at the Uganda Cancer Institute (UCI), a high proportion of patients with HIV-associated lymphoma attending UCI were initially misdiagnosed and treated as TB (40).

In our study, we investigated possible non-TB etiologies which could occur as comorbidities or as alternative diagnosis in this population of X-pert negative PLHIV. We found high frequency of cryptococcosis, and bacterial blood stream infections including Staphylococcus aureus, E. coli among others, which may have contributed to the high mortality. Among the investigations done were sputum TB culture and urine TB-LAM, and we found that 12 of the 67 (17.9%) non-treated patients that died had a positive TB culture and/or positive urine TB-LAM. It should be noted that these TB investigations were not routine at the time of the study, and thus these patients were considered non-TB by the attending clinician. This highlights the diagnostic challenges in detecting paucibacillary TB among PLHIV particularly with advanced disease, where even sensitive tests like Xpert have reduced sensitivity, and the need for additional diagnostic approaches including TB culture and TB-LAM in high HIV/TB prevalence settings.

Of the participants found with cryptococcosis, almost 90% were empirically treated for TB, yet more than 40% died, highlighting how misdiagnosis of opportunistic infections contributes to mortality in advanced HIV disease even in the presence of empiric TB treatment (21, 24, 25).

The loss to follow-up in this study was 7%, and although minimal, it could have introduced bias into the mortality estimates, particularly for the participants in whom empiric TB treatment was not considered. The absence of continuous monitoring and in-depth follow-up in some cases, further limits the generalizability of these findings. Other limitations include the lack of post-mortem data to confirm causes of death and the limited availability of comprehensive HIV monitoring testing including viral load data.

## Conclusion

In this TB/HIV high-burden setting, we found high mortality among Xpert-negative presumptive TB adult PLHIV, occurring mostly within the first three months post-enrolment and predominantly among hospitalized individuals. Mortality was higher among those empirically treated for TB compared to those who were not, even in this population where the majority were on ART.

While to some extent, empiric TB treatment may remain necessary for PLHIV with negative Xpert results, co-infections, particularly cryptococcemia and bacteremia, are not uncommon, underscoring the need to vigorously investigate for non-TB conditions in this patient population. Expanding access to diagnostics including TB culture, bacterial cultures, and tests for the common opportunistic infections and other HIV-associated disease conditions such as lymphoma, alongside adequate follow-up systems, is essential to mitigate mortality.

## Data Availability

All relevant data are within the manuscript and its Supporting Information files

## Acknowledgements

The authors would like to thank the research team particularly Bernadette Nakawooya, Raymond Serubiri, Joseph Kiwanuka, Betty Namboozo, Esther Iriamo, the Infectious Diseases Institute Datafax team and the Mycobacteriology (BSL-3) Laboratory of Makerere University for their important contributions to the implementation of this study. We thank the staff at Mulago National Referral Hospital and Kisenyi Health Centre-IV for their support throughout the study. The authors gratefully acknowledge the study participants for their willingness to participate in the study.

## Authors’ contribution

LN, YCM, HMK conceived the idea; LN, YCM obtained funding, LN, SN, LN, LK, BB, WS, BN acquired the data; LN, BK, AJ, WS, FK, YCM, HMK performed the analysis and interpretation, LN primarily drafted the article. All authors contributed to revising this manuscript and gave final approval of the version.

